# Shared Neurocardiac Pathways Linking Atrial Fibrillation and Depression: A UK Biobank Analysis

**DOI:** 10.64898/2026.02.21.26346796

**Authors:** Charles Verdonk, Aleksandr Talishinsky, Navid Hakimi, Masaya Misaki, Jonas L. Steinhäuser, Wesley K. Thompson, Chun Chieh Fan, Martin P. Paulus, Olujimi A. Ajijola, Sahib S. Khalsa

## Abstract

Central Illustration

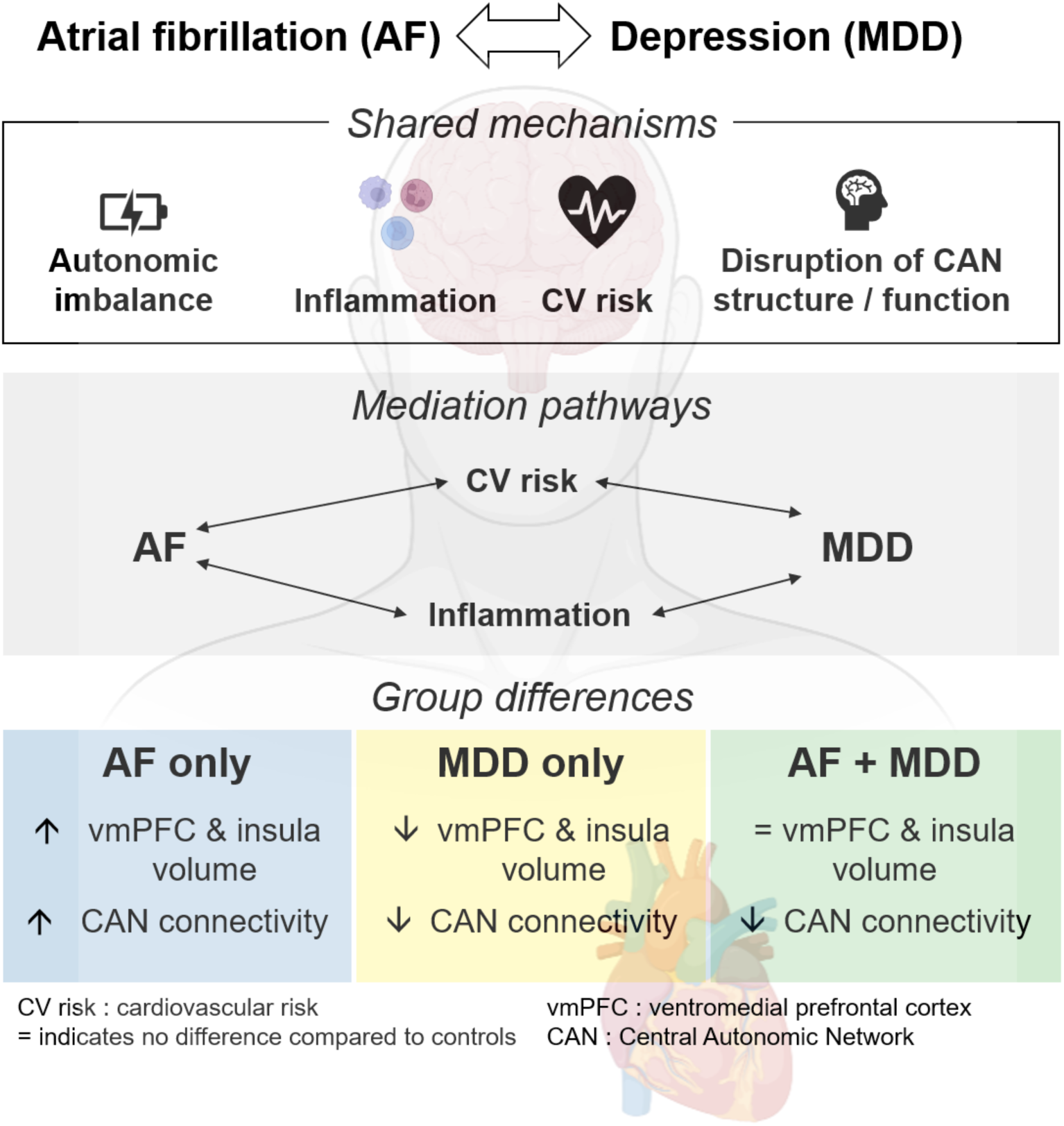

**HIGHLIGHTS:**

- Atrial fibrillation and depression are linked via central autonomic network disruption, cardiovascular risk, and inflammation.
- Heightened inflammatory response and cardiovascular risk mediates the bidirectional relationship between atrial fibrillation and depression.
- Atrial fibrillation, depression, and their comorbidity exhibit distinct, non-additive neural and autonomic signatures.

**BACKGROUND:** Atrial fibrillation (AF) and major depressive disorder (MDD) frequently co-occur and are each associated with adverse cardiovascular outcomes, yet the biological pathways linking these conditions remain poorly defined. Using the UK Biobank, we evaluated shared neurocardiac, inflammatory, and cardiovascular correlates underlying the AF-MDD association.

**OBJECTIVES:** To assess bidirectional associations between AF and MDD and determine whether shared inflammatory, cardiovascular, autonomic, and neuroimaging correlates characterize their comorbidity.

**METHODS:** We analyzed individuals with AF (N≥1,716), MDD (N≥4,550), comorbid AF-MDD (N≥243), and healthy comparators (HCs; N≥33,041). Bidirectional associations were examined using cross-sectional and Cox proportional hazard models. Mediation analyses evaluated contributions of inflammatory markers and cardiovascular risk. Central autonomic network structure and function was assessed using MRI-derived morphometry and resting-state connectivity.

**RESULTS:** AF and MDD demonstrated bidirectional associations: AF was associated with a 44% higher risk of incident MDD, and MDD with a 26% higher risk of incident AF. Inflammatory biomarkers and cardiovascular risk partially mediated these associations (6.85% and 32.01%, respectively). AF was associated with greater gray matter volume in ventromedial prefrontal and insular cortices and increased central autonomic network connectivity, whereas MDD showed opposite structural and functional patterns. The comorbid AF-MDD group exhibited distinct, non-additive neural profiles.

**CONCLUSIONS:** AF and MDD demonstrate bidirectional associations characterized by shared inflammatory, cardiovascular, and neural correlates, alongside distinct and non-additive alterations within central autonomic network circuits. These findings support a systems-level neurocardiac framework linking cardiac and psychiatric disease and highlight the importance of integrated approaches to risk assessment and multidisciplinary management in patients with AF-MDD comorbidity.

## INTRODUCTION

Atrial fibrillation (AF) is the most common sustained cardiac arrhythmia^1^ and is associated with substantial morbidity, mortality, and healthcare burden worldwide.^2,3^ Its prevalence has increased markedly over recent decades, reflecting aging populations and rising cardiometabolic risk factors.^4,5^ Major depressive disorder (MDD) is similarly common and is independently associated adverse cardiovascular outcomes, including increased risk of incident cardiovascular disease and mortality.^6^ Together, AF and MDD represent highly prevalent conditions that frequently co-occur,^7,8^ and contribute significantly to global disease burden.

Epidemiologic studies consistently demonstrate a bidirectional relationship between AF and depression. MDD affects approximately 24.3% of adults with AF (and up to 40.3% in older adults),^9–11^ and is associated with worse clinical outcomes, reduced quality of life, and increased healthcare utilization.^9,12^ Conversely, depression is associated with an increased risk of incident AF, with meta-analyses and large population cohorts reporting 25% higher AF risk among individuals with MDD.^13,14^ Genetic and longitudinal studies further suggest potential causal links, via a positive association between the frequency of depression symptoms and AF incidence (with hazard ratios (HR) ranging from 1.25 to 2.17),^15^ and evidence from Mendelian randomization studies supporting a causal relationship from MDD to AF,^16,17^ although the biological pathways underlying this association remain poorly understood. Elucidating shared mechanisms linking AF and MDD is therefore critical for improving risk stratification and developing integrated treatment approaches.

Several overlapping pathophysiological processes have been implicated in both conditions. Autonomic imbalance, characterized by increased sympathetic and reduced parasympathetic activity, plays a central role in AF initiation and maintenance^18^ and is also a well-established feature of depression.^19^ Similarly, systemic inflammation is increasingly recognized as a key contributor to both AF pathogenesis and depressive illness, with elevated pro-inflammatory cytokines commonly observed in both conditions.^20–22^ Poor cardiovascular health and cardiometabolic risk factors further contribute to the development of AF^23^ and are strongly associated with depression,^19^ suggesting shared systemic pathways.

Emerging evidence also highlights the importance of central neurocardiac mechanisms in both conditions. Neuroimaging studies in MDD consistently demonstrate structural and functional alterations in brain regions involved in autonomic regulation, including the insular cortex and medial prefrontal cortex.^24–27^ Functionally, disrupted connectivity in specific brain networks has been proposed to play a mediating role in depression, including the default mode network (DMN), the central executive network (CEN), and the salience network (SN).^24,28^ In addition, dysfunction of the central autonomic network (CAN),^29^ which plays a pivotal role in regulating peripheral autonomic function, has also been proposed such that CAN activity may even mediate the link between inflammatory processes and autonomic imbalance in depression.^30,31^ These findings support the role of the CAN as a potential integrative substrate linking cardiac and psychiatric pathology. Notably, multimodal imaging work has further demonstrated associations between AF and alterations in brain structure and connectivity within autonomic regulatory regions.^32–34^ Lesions in the right insula have even been associated with the onset of AF^35^, with cardiac effects thought to be mediated by autonomic dysregulation, specifically an upregulation of sympathetic activity.^36^ Supporting this notion, abnormal heartbeat-evoked neural responses have implicated the right insular cortex in AF.^37^ Functional neuroimaging studies using resting-state MRI have further shown disrupted connectivity within the DMN in stroke-free AF patients^33,37,38^ Despite these advances, no large-scale study has comprehensively examined whether shared peripheral and central mechanisms jointly characterize AF-MDD comorbidity. In particular, it remains unclear whether inflammatory, cardiovascular, autonomic, and neuroimaging correlates converge to explain the bidirectional association between these conditions.

In the present study, we leveraged the UK Biobank^39–41^ to test the hypothesis that shared neurocardiac pathways spanning peripheral biological systems and central autonomic network alterations link AF and MDD. We examined bidirectional associations between AF and MDD and evaluated whether inflammatory markers, cardiovascular risk, autonomic function, and structural and functional brain measures within key CAN regions jointly characterize their comorbidity.

We hypothesized that the association between AF and MDD is mediated by shared mechanisms across four domains: autonomic imbalance, heightened inflammatory response, poor cardiovascular health, and structural and functional brain alterations within the CAN. Our analysis specifically focused on two cortical regions consistently implicated in CAN activity—the ventromedial prefrontal cortex (vmPFC) and the insular cortex.^42^ We hypothesized that, when examined independently, both AF and MDD would be associated with: (1) autonomic imbalance characterized by parasympathetic underactivity; (2) a heightened inflammatory response marked by elevated pro-inflammatory cytokines; and (3) poor cardiovascular health reflected in high cardiovascular risk profiles. At the brain level, we predicted alterations in CAN structure and function, including reduced gray matter volume in the vmPFC and insula, as well as diminished network connectivity within the CAN and between the CAN and other major large-scale brain networks, such as the DMN, CEN, and SN. Furthermore, we anticipated that individuals with comorbid AF and MDD would show greater disruption across both peripheral biological systems (autonomic, inflammatory, and cardiovascular markers) and central nervous system indicators (structural and functional changes in the CAN) compared to those with either condition alone.

## METHODS

### Study Population

Between 2006 and 2010, the UK Biobank enrolled 503,325 individuals aged 40 to 69 years who were registered with general practitioners under the UK National Health Service.^41^ At recruitment, detailed information on participants’ demographics, medication use, and health outcomes was collected through questionnaires and medical records. Baseline blood samples were also taken to analyze serum biomarkers, including those identified as established disease risk factors or commonly assessed in clinical evaluations. Neuroimaging data were acquired as well, covering structural, diffusion, and resting-state functional imaging modalities.^43^ Additionally, participants underwent an electrocardiogram (ECG) test, which included a 15-second resting phase (protocol details are available online: https://biobank.ndph.ox.ac.uk/showcase/ukb/docs/Cardio.pdf). Informed consent was obtained from all participants, and the study protocol received approval from the North West Multicenter Research Ethics Committee. The study adhered to the principles outlined in the Declaration of Helsinki.

### Definition of Cases (Atrial fibrillation, Depression) and Controls

Atrial fibrillation (AF) diagnosis was determined based on ICD-10 code I48, which includes both AF and atrial flutter (UK Biobank field: 131350). Because ICD-10 I48 does not distinguish paroxysmal from persistent AF, the AF category reflects a heterogeneous mix of arrhythmia subtypes. The identification of Major Depressive Disorder (MDD) cases relied on multiple data sources.^44^ Depression diagnoses, either primary or secondary, were identified through a self-reported question (UK Biobank field: 20002, values 1286 and 1531) and linked hospital admission records, utilizing the following ICD-10 codes: F32 (single-episode depression; UK Biobank field: 130894), F33 (recurrent depression; UK Biobank field: 130896), F34 (persistent mood disorders; UK Biobank field: 130898), and F38 (other mood disorders; UK Biobank field: 130900). At enrollment, participants also completed the 4-item Patient Health Questionnaire (PHQ-4), which comprises a 2-item depression scale (PHQ-2) and a 2-item Generalized Anxiety Disorder scale.^45^ MDD caseness was defined as a positive response to the self-reported question, a clinical diagnosis of depression, or a PHQ-2 score of 3 or higher (the cutoff for depressive disorder^46^) at the baseline assessment. Because PHQ-2 scores may capture milder or subclinical depressive symptoms, the resulting MDD group likely includes individuals with a broader range of severity than typical clinical cohorts. As a control group, population controls were identified as UK Biobank participants who had no recorded diagnosis of AF or MDD. This approach resulted in datasets with varying numbers of cases and controls depending on the data available for each data type (e.g., cardiac data and neuroimaging data; Table S1 in Supplementary Methods 1). Accordingly, both AF and MDD case definitions reflect clinically relevant but heterogeneous groups, consistent with prior population-based analyses in the UK Biobank.

### Data processing

Except for connectivity and graph theory metrics derived from resting-state functional imaging, we used the measures generated by UK Biobank (https://www.ukbiobank.ac.uk/).

### Connectivity and graph theory metrics derived from MRI data

The present study utilized resting-state fMRI data from the initial scanning visit of the UK Biobank. The preprocessed resting-state fMRI data^47^ were normalized to MNI template space using the provided warp parameter files. The mean signal time course was extracted for each of the 246 regions defined in the Brainnetome atlas.^48^ A bandpass filter (0.009–0.08 Hz) was applied, and time points with a frame-wise displacement (FD) greater than 0.3 mm were censored. Pearson correlation coefficients were computed between regional time courses and subsequently z-transformed to obtain functional connectivity (FC).

We focused on four large-scale brain networks: the central autonomic network (CAN), central executive network (CEN), salience network (SN), and default mode network (DMN). The Brainnetome atlas regions corresponding to each network are listed in Table S2 (Supplementary Methods 2). Within-network FC was calculated as the average FC between regions within the same network, while between-network FC was computed as the average FC across all pairwise connections between individual regions from two different networks. Because these network metrics are highly correlated, we applied a unified Holm correction across all network-level comparisons rather than treating each metric independently.

Additionally, we assessed network metrics using a graph theory approach.^49^ These metrics were derived from a weighted graph of positive FC using the NetworkX library in Python (https://networkx.org/documentation/stable/index.html).^50^ The network metrics included global efficiency,^51^ clustering coefficient,^52^ modularity,^53^ betweenness centrality,^54^ and eigenvector centrality.^55^ The node-specific values were averaged across nodes within each network. Global efficiency is the average inverse shortest path length between nodes, measuring how efficiently information travels within the network. The clustering coefficient quantifies the likelihood that a node’s neighbors are also connected, reflecting local interconnectedness. Modularity measures the extent to which a network is divided into distinct modules. Betweenness centrality is the sum of the fractions of all-pairs shortest paths that pass through a given node, indicating how often a node serves as a bridge between other nodes. Eigenvector centrality measures a node’s importance based on the centrality of its connections, with higher values identifying key hub regions.

### Cardiovascular risk profile

To define the cardiovascular risk profile, we adopted a previously published definition of metabolic syndrome from UK Biobank data, which has been shown to mediate the relationship between arterial stiffness and major depression.^44^ A ‘high’ cardiovascular risk profile (i.e. ‘metabolic syndrome’) was defined as the presence of three or more of the following: unhealthy waist circumference, hypertension, dyslipidemia, and hypertriglyceridemia (Supplementary Methods 3).

### Inflammatory response

We used continuous data from the UK Biobank database to assess circulating levels of inflammatory acute-phase and immune-related markers in participant serum, including C-reactive protein (CRP; UK Biobank field: 30710), insulin-like growth factor (IGF-1; UK Biobank field: 30770), and rheumatoid factor (RF; UK Biobank field: 30820). While major depressive disorder (MDD) has been associated with elevated levels of multiple proinflammatory cytokines,^20^ these canonical cytokines are not available as blood-based measures in the UK Biobank dataset.

### Cardiac data

To complement these data, exploratory analyses of heart rate variability (HRV) metrics derived from resting electrocardiogram (ECG) recordings were conducted. Ultra-short HRV (usHRV) metrics were extracted from 15-second resting ECG recordings using a processing pipeline implemented in MATLAB 2020b (MathWorks®; Supplementary Methods 4). The HRVAS-master toolbox (version 1.0.2) was used to compute the Root Mean Square of Successive Differences (RMSSD) and the Standard Deviation of Successive Differences (SDSD).^56,57^ High-frequency power (PHF) in the 0.15–0.40 Hz spectral band was computed using scripts developed by Candia-Rivera.^58^ These usHRV metrics from the UK Biobank dataset have been previously validated through their strong agreement with standard HRV measures obtained from 6-minute ECG recordings.^56^

### Analysis strategy

#### Mediation analysis

To explore the relationship between AF and MDD, we quantified both indirect associations mediated through inflammatory markers and cardiovascular risk profile, as well as direct associations independent of these mediators. Three separate multivariate logistic regression models were employed: one to evaluate the total effect (the overall relationship between AF and MDD without accounting for mediation), another to assess the mediated effect (the association between AF and the mediators), and a third to determine the direct effects (the relationships between AF and MDD after accounting for the mediation pathway, as well as between the mediators and MDD). The statistical significance of mediation effects was tested using a bootstrapping approach with 500 replications to ensure accurate estimation of standard errors for indirect effects.^59^ For significant mediation effects, the proportion of the total effect mediated was calculated to quantify the magnitude of mediation (Supplementary Methods 5. Mediation analysis).^60^

### Cox proportional hazard models

Incident MDD diagnosis was defined as the time interval (in years) between the date of a positive AF diagnosis (UK Biobank field: 131350) and the date of a positive MDD diagnosis (UK Biobank fields: 130894, 130896, 130898, and 130900) for participants categorized as AF+/MDD+. The control group included participants without an AF diagnosis, classified as AF-/MDD-(without MDD) or AF-/MDD+ (with MDD). For control participants who developed MDD (AF-/MDD+), the follow-up starting point was matched to the date of AF diagnosis from age-matched participants in the AF+/MDD+ group. Data from participants without an MDD diagnosis were censored on August 1, 2022, the date of the most recent recorded MDD diagnosis (Figure S3A in Supplementary Methods 6).

Conversely, incident AF diagnosis was defined as the time elapsed (in years) from the date of a positive MDD diagnosis to the date of a positive AF diagnosis for participants categorized as MDD+/AF+. The control group included individuals without an MDD diagnosis, further divided into MDD-/AF-(neither MDD nor AF) and MDD-/AF+ (AF without MDD). For control participants who developed AF (MDD-/AF+), the follow-up began at a point equivalent to the date of MDD diagnosis in age-matched MDD+/AF+ participants. Participants without an AF diagnosis were censored on July 1, 2022, marking the most recent recorded AF diagnosis (Figure S3B in Supplementary Methods 6).

Cox proportional hazards models were used for time-to-event analyses to evaluate the association between AF and the risk of developing MDD, and vice versa, while adjusting for age and sex.

### Association analysis

Descriptive statistics (e.g., mean, standard deviation (SD)) and multivariate linear regression were used to compare group differences among cases (AF, MDD, or comorbid AF-MDD) and healthy controls (HCs) for variables not included in the mediation analyses. The neuroimaging measures were excluded from mediation analysis because they originated from datasets with smaller sample sizes, in which the association between AF and MDD did not reach statistical significance, a necessary condition for conducting mediation analysis. To account for multiple comparisons in association analysis, the Holm correction was applied to adjust p-values and control the family-wise error rate. The Holm approach was selected because it provides strong control of the family-wise error rate while maintaining greater power than Bonferroni correction in correlated neuroimaging and physiological measures.

### Covariates

Factors associated with both AF and MDD, including self-reported age and sex (female vs. male), were included as confounders in all analyses.^15,61–64^ Indicators of socioeconomic status and educational attainment - both of which have been associated with AF^65^ and with depression^66^ - were additionally included in association analyses; however, as their inclusion did not materially alter the strength or direction of the AF-MDD associations, they were not retained as covariates in subsequent analyses.

Statistical analyses were conducted using R version 4.2.2, with the following packages employed for specific purposes: ‘MASS’ for calculating 95% confidence intervals (CI) of model parameters^67^ ‘effectsize’ for estimating effect sizes,^68^ ‘boot’ for applying the bootstrapping approach to assess the statistical significance of mediation effects,^69^ and ‘survival’ for performing survival analyses using Cox proportional hazard models.^70^

### Data Sharing Statement

Individual-level data used in the present study are available upon application to the UK Biobank (https://www.ukbiobank.ac.uk). All other relevant data are available upon reasonable request to the authors.

## RESULTS

### Descriptive statistics of the analyzed sample

Among 502,180 UK Biobank participants with analyzable data, 38,041 (7.56%) had a diagnosis of Atrial Fibrillation (AF), and 80,640 (16.06%) met criteria for Major Depressive Disorder (MDD). Comorbid AF and MDD were observed in 6,869 participants, representing 1.37% of the total sample. The mean age of the total sample was 72.46 years (standard deviation (SD) = 8.12), and 54.4% of participants were female.

### Association between Atrial fibrillation and Major depressive disorder

AF was significantly and bidirectionally associated with MDD: patients with AF were more likely to have MDD than those without AF (β=0.31; p<0.001; OR=1.37, 95% CI [1.33; 1.41]), and patients with MDD were more likely to have AF than those without MDD (β=0.35; p<0.001; OR=1.41, 95% CI [1.37; 1.45]). These associations remained robust across sensitivity analyses using alternative PHQ-2–based definitions of depressive symptom severity, including both low- and high-frequency symptom endorsements (Supplementary Results 1).

### Survival analysis

AF condition was associated with a significantly increased risk of developing MDD (hazard ratio (HR)=1.44; 95% CI: [1.36;1.52]; p<0.001), with a shorter mean time to MDD diagnosis compared to individuals without AF (6.39 vs. 8.96 years Figure 1A). Conversely, individuals with MDD had a higher risk of developing AF (HR=1.26; 95% CI: [1.22;1.30]; p<0.001), with a shorter mean time to AF diagnosis than those without MDD (16.8 vs. 18.5 years; Figure 1B).

**Figure 1.**
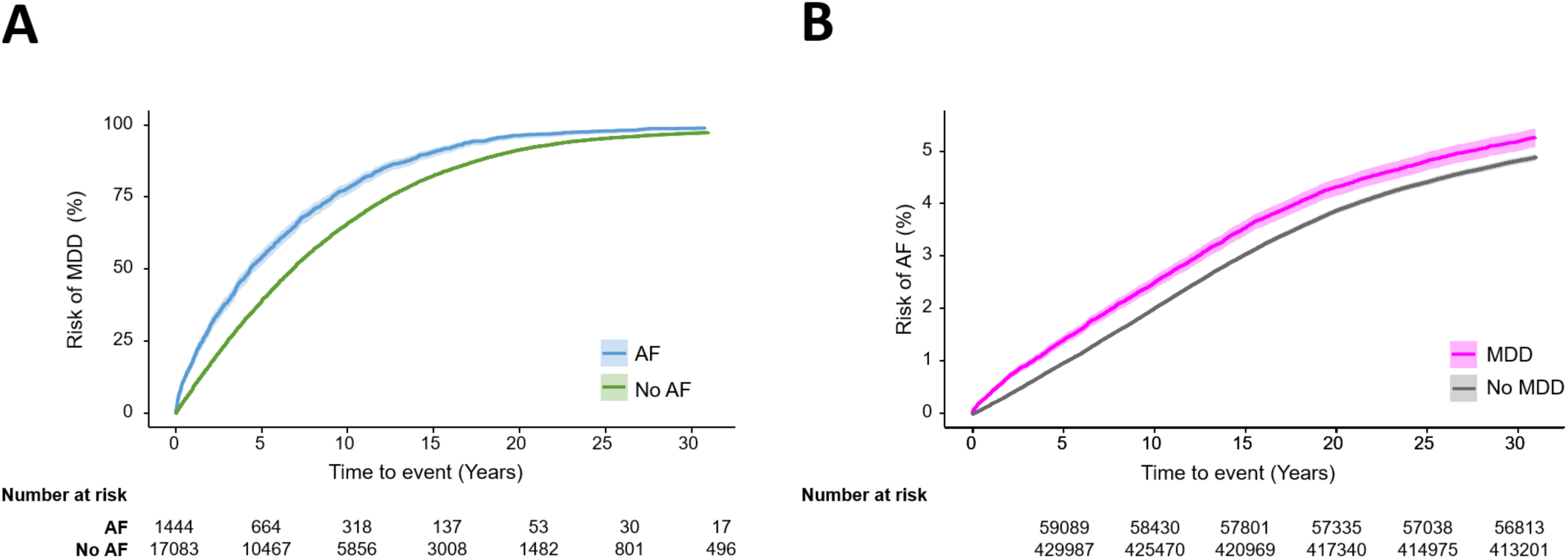
Survival analysis findings. **(A)** Atrial Fibrillation (AF) condition was associated with a significantly higher risk of Major Depressive Disorder (MDD) over time. **(B)** Individuals with MDD had a significantly higher risk of developing AF during follow-up.

### Mediation analysis

A high cardiovascular risk profile mediated 32.01% of the association between AF and MDD (95% CI [30.54;33.71]; Figure 2A; Table S3 in Supplementary Results 2).

**Figure 2.**
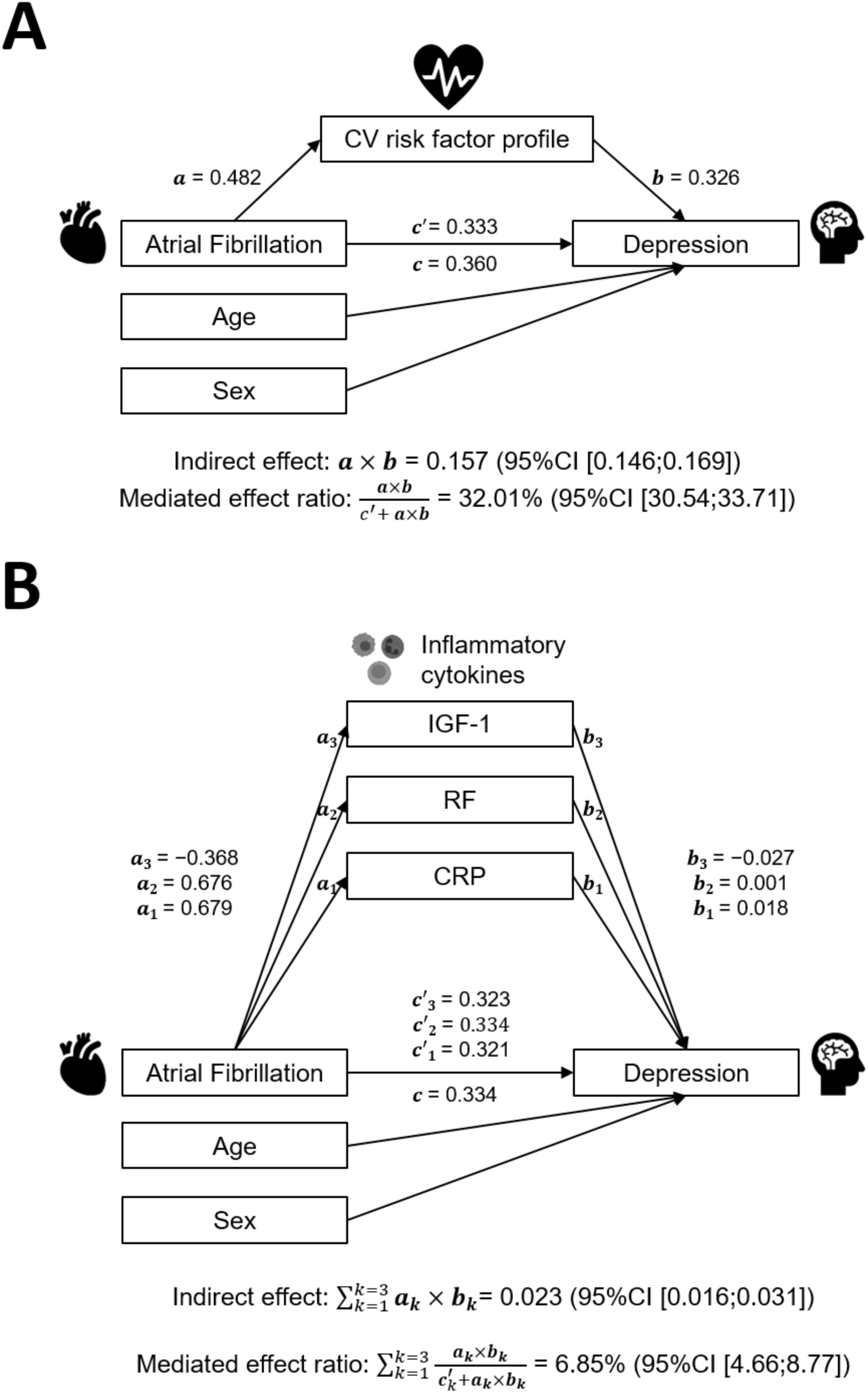
Mediation findings. A high cardiovascular (CV) risk profile **(A)** and a heightened inflammatory response **(B)** mediated the positive association between Atrial Fibrillation and Depression. The heightened inflammatory response was indexed by increased C-reactive protein (CRP) and rheumatoid factor (RF) levels and decreased insulin-like growth factor (IGF-1) levels.

A heightened inflammatory response, reflected by increased serum levels of C-reactive protein (CRP) and rheumatoid factor (RF), and lower serum levels of insulin-like growth factor (IGF-1) mediated 6.85% of the association between AF and MDD (95%CI [4.66;8.77]; Figure 2B; Table S3 in Supplementary Results 2).

### Association analysis

#### Neuroimaging findings

Significant group comparisons for neuroimaging outcomes are summarized in Table 1, Figure 3 and Figure 4.

**Figure 3.**
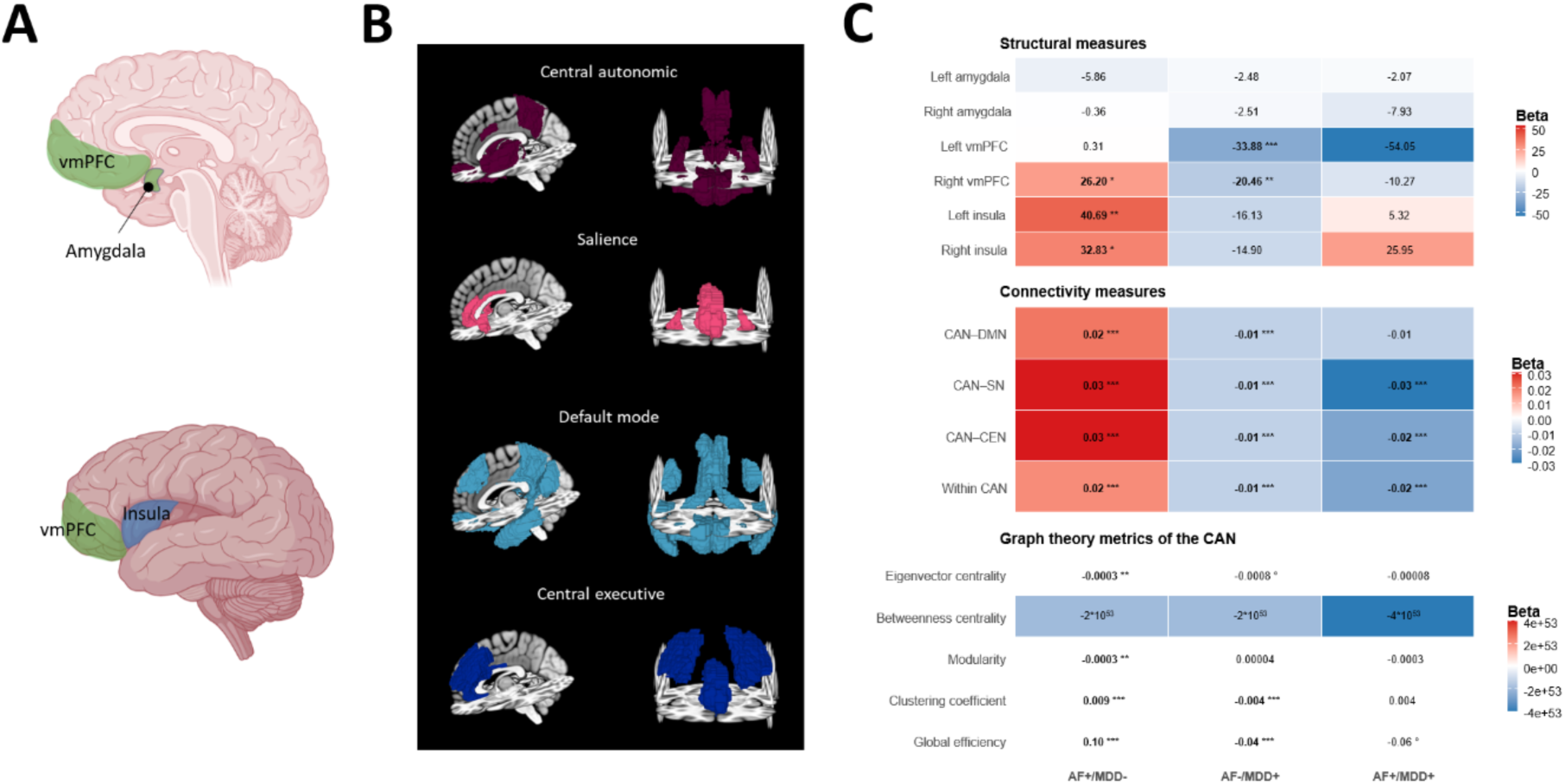
Neuroimaging correlates of atrial fibrillation and major depressive disorder. **(A)** Brain regions of interest included in structural analyses (insula, ventromedial prefrontal cortex, and amygdala). **(B)** Intrinsic brain networks examined in functional connectivity and graph-theoretical analyses. **(C)** Heatmap summarizing beta coefficients from group-wise association analyses of structural, functional connectivity, and graph-theoretical measures, comparing individuals with atrial fibrillation only (AF+/MDD−), major depressive disorder only (AF−/MDD+), and comorbid AF–MDD (AF+/MDD+), relative to healthy controls. Warm colors indicate positive associations (increases), and cool colors indicate negative associations (decreases). Asterisks denote statistically significant group differences (\**p* < 0.05, \*\**p* < 0.01, \*\*\**p* < 0.001), while ° indicates trend-level significance. Measures are organized by domain, including regional gray matter volume (upper pannel), connectivity (middle panel), and CAN-related graph-theoretical metrics (bottom pannel). Abbreviations: vmPFC, ventromedial prefrontal cortex; INS, Insula; DMN, Default Mode Network; CEN, Central Executive Network; SN, Salience Network; CAN, Central Autonomic Network; GEff, Global Efficacy; ClustCoef, Clustering Coefficients; Modu, Modularity; EigCent, Eigenvector Centrality; BetwCent, Betweenness Centrality.

**Figure 4.**
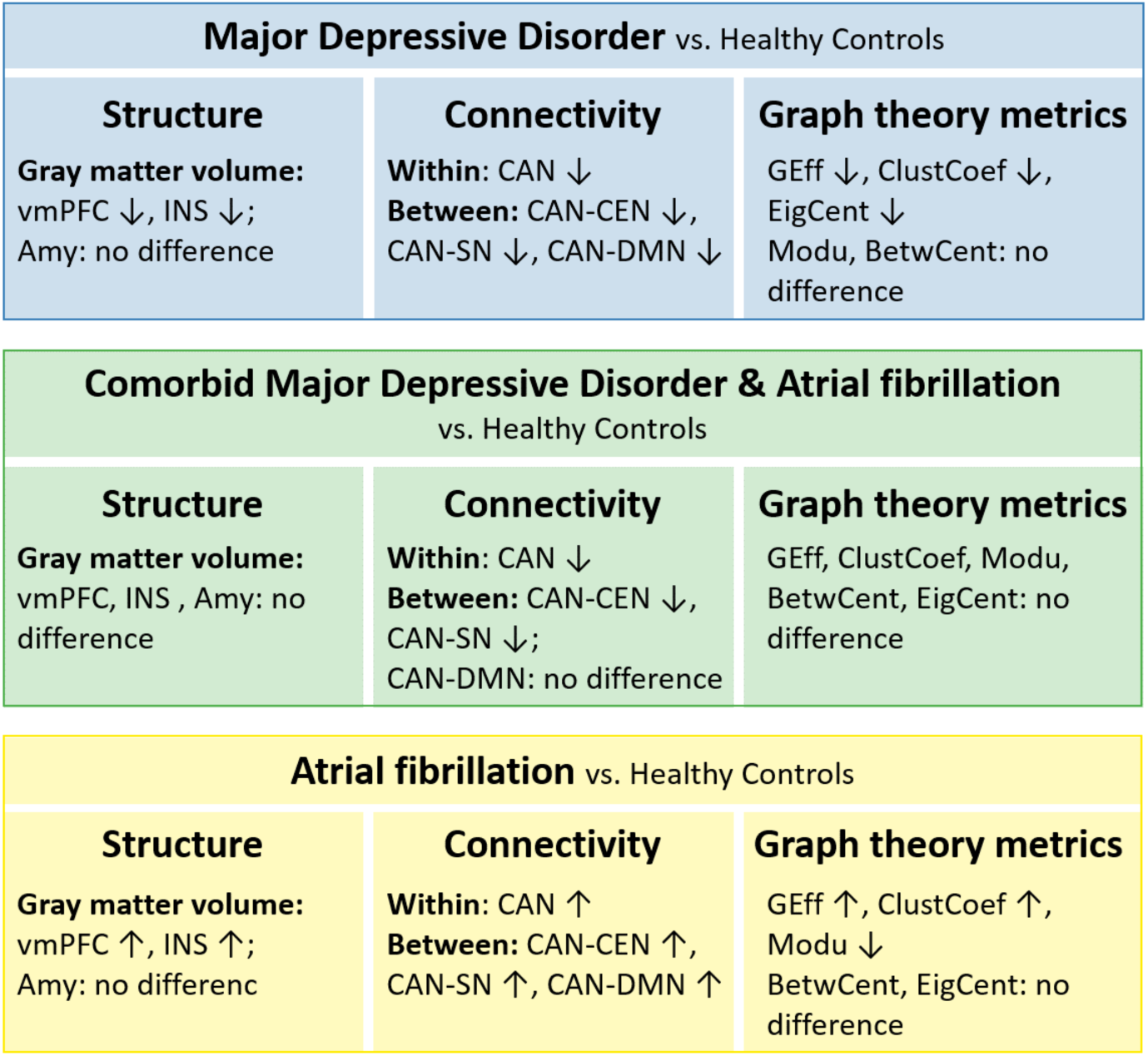
Graphical summary of neuroimaging association in atrial fibrillation and major depressive disorder. Group-level differences in structural and functional neuroimaging outcomes, comparing individuals with atrial fibrillation (AF), major depressive disorder (MDD), or both, to healthy controls (HCs). Abbreviations: vmPFC, ventromedial prefrontal cortex; INS, Insula; Amy, Amygdala; DMN, Default Mode Network; CEN, Central Executive Network; SN, Salience Network; CAN, Central Autonomic Network; GEff, Global Efficacy; ClustCoef, Clustering Coefficients; Modu, Modularity; EigCent, Eigenvector Centrality; BetwCent, Betweenness Centrality.

**Table 1.** Summary of statistical results from group comparisons of neuroimaging measures. Results of association analyses across patient groups with atrial fibrillation (AF), major depressive disorder (MDD), and comorbid AF–MDD, relative to a reference group of healthy controls (HCs).

| *Outcome* | *Group*<br>*N (%)* |  |  |  |
| --- | --- | --- | --- | --- |
| **Structural** | HC (Reference)<br>35,737 (82.89) | AF+/MDD-<br>1,820 (4.22) | AF-/MDD+<br>5,342 (12.39) | AF+/MDD+<br>221 (0.51) |
| Right insula | - | ↑* | NS | NS |
| Left insula | - | ↑** | ↓° | NS |
| Right vmPFC | - | ↑* | ↓** | NS |
| Left vmPFC | - | NS | ↓*** | NS |
| Right amygdala | - | NS | NS | NS |
| Left amygdala | - | NS | NS | NS |
| **Connectivity** | HC (Reference)<br>33,948 (81.41) | AF+/MDD-<br>1,716 (4.12) | AF-/MDD+<br>5,793 (13.89) | AF+/MDD+<br>243 (0.58) |
| Within CAN | - | ↑*** | ↓*** | ↓* |
| CAN-CEN | - | ↑*** | ↓*** | ↓° |
| CAN-SN | - | ↑*** | ↓*** | ↓* |
| CAN-DMN | - | ↑*** | ↓*** | NS |
| **Graph theory (CAN)** | HC (Reference)<br>33,948 (81.41) | AF+/MDD-<br>1,716 (4.12) | AF-/MDD+<br>5,793 (13.89) | AF+/MDD+<br>243 (0.58) |
| Global Efficacy | - | ↑*** | ↓*** | ↓° |
| Clustering Coefficients | - | ↑*** | ↓*** | NS |
| Modularity | - | ↓** | NS | NS |
| Betweenness Centrality | - | NS | NS | NS |
| Eigenvector Centrality | - | NS | ↓** | ↓° |

NS: No significance difference with the reference group (HCs); ↑: higher value than the reference group (HCs); ↓: lower value than the reference group (HCs); Statistical significance after Holm correction: \* $p < 0.05$ ; \*\* $p < 0.01$ ; \*\*\* $p < 0.001$ ; ° $p < 0.10$ ; NS: non-significant; vmPFC: ventromedial prefrontal cortex; PTR: Posterior thalamic radiations; CAN: Central Autonomic Network; CEN: Central Executive Network;

### Structural data (Gray matter volume)

Compared to HCs, individuals in the AF+/MDD− group showed a significantly larger gray matter volume in the right insula (β=32.83, p=0.03). No significant differences were observed in the right insula for the AF−/MDD+ (β=−14.90, p=0.11) or AF+/MDD+ (β=25.95, p=0.54) groups. In the left insula, a significant volume increase was also found in the AF+/MDD− group (β=40.69, p=0.006), while the AF−/MDD+ group showed a trend toward reduced volume (β=−16.13, p=0.07). No significant difference was detected in the AF+/MDD+ group (β=5.32, p=0.90).

Gray matter volume in the right ventromedial prefrontal cortex (vmPFC) was significantly larger in the AF+/MDD-group (β=26.20, p=0.04) and significantly smaller in the AF-/MDD+ group (β=-20.46, p=0.009) relative to HCs. No significant difference was observed in the AF+/MDD+ group (β=−10.27, p=0.77). For the left vmPFC, only the AF−/MDD+ group showed a significant volume reduction compared to HCs (β=−33.88, p<0.001), whereas the AF+/MDD− (β=0.31, p=0.98) and AF+/MDD+ (β=−54.05, p=0.15) groups did not differ significantly.

For the amygdala, no significant differences were found across groups. In the left amygdala, gray matter volumes were comparable to controls in the AF+/MDD− (β=−5.86, p=0.22), AF−/MDD+ (β=−2.48, p=0.39), and AF+/MDD+ (β=−2.07, p=0.87) groups. The same pattern was observed in the right amygdala (AF+/MDD−: β=−0.36, p=0.94; AF−/MDD+: β=−2.51, p=0.39; AF+/MDD+: β=−7.93, p=0.55).

### Connectivity measures (Resting state functional connectivity)

Compared to HCs, connectivity within the Central Autonomic Network (CAN) was significantly increased in the AF+/MDD-group (β=0.02, p<0.001), but decreased in both the AF-/MDD+ (β=-0.01, p<0.001) and the AF+/MDD+ (β=-0.02, p<0.001) groups.

Connectivity between the CAN and the Central Executive Network (CEN) followed a similar pattern: it was higher in the AF+/MDD-group (β=0.03, p<0.001), but lower in the AF-/MDD+ (β=-0.01, p<0.001) and the AF+/MDD+ (β=-0.02, p<0.001) groups.

For CAN–Salience Network (SN) connectivity, we also observed increased connectivity in the AF+/MDD-group (β=0.03, p<0.001), and reduced connectivity in the AF-/MDD+ (β=-0.01, p<0.001) and the AF+/MDD+(β=-0.03, p<0.001) group.

Finally, connectivity between the CAN and the Default Mode Network (DMN) was higher in the AF+/MDD-group (β=0.02, p<0.001), but lower in the AF-/MDD+ group (β=-0.01, p<0.001). No significant difference was observed in the AF+/MDD+ group (β=-0.01, p=0.22).

### Graph theory metrics of the Central Autonomic Network (CAN)

Compared to HCs, global efficiency was significantly higher in the AF+/MDD− group (β=0.10, p<0.001), significantly lower in the AF−/MDD+ group (β=-0.04, p<0.001), and showed a trend toward reduction in the AF+/MDD+ group (β=-0.06, p=0.08). Clustering coefficient values were significantly elevated in the AF+/MDD− group (β=0.009, p<0.001), significantly reduced in the AF−/MDD+ group (β=-0.004, p<0.001), and not significantly different in the AF+/MDD+ group (β=0.004, p=0.29). Regarding modularity, only the AF+/MDD− group showed significantly lower values compared to HCs (β=-0.0003, p=0.002), while no significant differences were observed for the AF−/MDD+ (β=0.00004, p=0.46) and AF+/MDD+ (β=-0.0003, p=0.33) groups. Eigenvector centrality was significantly reduced in the AF−/MDD+ group (β=-0.0003, p=0.006), showed a trend toward reduction in the AF+/MDD+ group (β=-0.0008, p=0.08), and did not differ significantly from HCs in the AF+/MDD− group (β=-0.00008, p=0.59). No significant group differences were found for betweenness centrality.

### Cardiac findings

Group comparisons for ultra-short heart rate variability (usHRV) outcomes are summarized in Table 2. Compared to healthy comparisons (HCs), the Root Mean Square of Successive Differences (RMSSD) values were significantly higher in the AF+/MDD-group (β=70.79, p<0.001) and the AF+/MDD+ group (β=37.97, p<0.001), but not in the AF-/MDD+ group (β=-2.14, p=0.28). A similar pattern was observed for the Standard Deviation of Successive Differences (SDSD), with significantly higher values in the AF+/MDD-(β=70.69, p<0.001) and AF+/MDD+ (β=40.23, p<0.001) groups, but no significant difference in the AF-/MDD+ group (β=-1.98, p = 0.33). For High-Frequency Power in the 0.15–0.40 Hz spectral band (PHF), only the AF+/MDD-group exhibited significantly elevated values relative to HCs (β=42.47, p<0.001). The AF-/MDD+ (β=3.66, p=0.26) and AF+/MDD+ (β=14.53, p=0.32) groups showed no significant differences.

**Table 2.** Summary of statistical results from group comparisons of heart rate variability measures. Results of association analyses across patient groups with atrial fibrillation (AF), major depressive disorder (MDD), and comorbid AF–MDD, relative to a reference group of healthy controls (HCs).

| Outcome | Group<br>N (%) |  |  |  |
| --- | --- | --- | --- | --- |
| Cardiac (usHRV) | HC (Reference)<br>38,972 (83) | AF+/MDD-<br>1,943 (4.14) | AF-/MDD+<br>5,793 (12.34) | AF+/MDD+<br>243 (0.52) |
| RMSSD | - | ↑*** | NS | ↑*** |
| SDSD | - | ↑*** | NS | ↑*** |
| PHF | - | ↑*** | NS | NS |

usHRV: ultra-short heart rate variability; RMSSD: Root Mean Square of Successive Differences; SDSD: Standard Deviation of Successive Differences; PHF: High-frequency power in the 0.15–0.40 Hz spectral band; NS: No significance difference with the reference group (HCs); ↑: higher value than the reference group (HCs); ↓: lower value than the reference group (HCs); Statistical significance after Holm correction: \*\*\* $p<0.001$ ; NS: non-significant;

## DISCUSSION

We observed robust bidirectional relationships between atrial fibrillation (AF) and major depressive disorder (MDD) across cross-sectional and time-to-event analyses, with partial statistical accounting by cardiovascular risk and inflammatory biomarkers. These findings extend prior epidemiologic evidence by demonstrating that AF and depression are not merely co-occurring conditions but represent dynamically linked disease processes that evolve over time. Neuroimaging analyses further differentiated these conditions at the level of central autonomic network (CAN) circuitry: AF was associated with greater ventromedial prefrontal and insular volumes and increased CAN connectivity, whereas MDD showed opposite structural and functional patterns. Individuals with comorbid AF–MDD exhibited distinct, non-additive neural profiles, suggesting that comorbidity reflects a unique neurocardiac phenotype rather than a simple additive disease burden. Exploratory ultra-short heart rate variability analyses suggested differential peripheral autonomic signatures, with increased parasympathetic-related variability in AF and comorbid AF–MDD, unlike MDD alone. Collectively, these results support a systems-level framework in which AF and MDD are linked through interacting cardiovascular, inflammatory, autonomic, and central neural pathways along the brain–heart axis.

Our findings build upon prior literature demonstrating an association between AF and depression. A recent meta-analysis reported approximately a 25% increased risk of AF among individuals with MDD,^14^ whereas our analysis demonstrated a stronger association, with a 41% higher odds of AF among individuals with MDD. Importantly, sensitivity analyses indicated that this association persisted when depression was defined using stricter or symptom-frequency–based PHQ-2 criteria, although the magnitude of the association was attenuated for more severe depressive symptoms. One possible explanation is the older age of participants in our sample (mean age: 72.46 years), compared to the younger cohorts (mean age 47–62) included in the aforementioned meta-analysis.^14^ Since AF incidence increases dramatically with age—rising nearly 15-fold between ages 40–49 and ≥70 years^64^—age may amplify the observed association. However, a large-scale South Korean cohort study (N>5 million) found that depression was more strongly associated with new-onset AF in younger adults and women,^13^ suggesting there may be complex age-related and sex-related interactions that warrant further investigation. The sex distribution in our study and the previously published meta-analysis^14^ was comparable, indicating sex is unlikely to explain the risk difference. Our findings are also consistent with prior AF studies reporting that approximately 38% of patients with AF meet criteria for depression,^9,10^ with prevalence rising to as high as 40.3% in older adults.^11^ Lastly, the observed 1.37% AF–MDD comorbidity in our study is consistent with the 1.6% reported by Kim et al. (2022) in their large-scale cohort study in South Korea.^13^ This slight difference may reflect age-related patterns in comorbidity prevalence across cohorts, although the small absolute magnitude also suggests that it is likely within the range of expected sampling variability.

In this study, we used Cox proportional hazards models to explore the temporal dynamics of the bidirectional association between AF and MDD. We found that individuals with AF had a 44% higher risk of subsequently developing MDD (hazard ratio of 1.44), while those with MDD had a 26% increased risk of incident AF (based on a hazard ratio of 1.26). To our knowledge, this is the first study to demonstrate such bidirectional risk in a large, well-characterized cohort with long-term follow-up data. Although Cox models establish temporal ordering, they do not demonstrate causality or bidirectional causal loops. However, Cox models offer a key advantage over simple regression techniques by incorporating time-to-event data, allowing us to estimate not only whether an association exists, but also when one condition tends to develop in relation to the other. Our finding that AF increases the risk of MDD strengthens prior reports based on smaller cohorts^9^ and supports a temporal role of AF in the onset of depression. Similarly, our findings align with recent UK Biobank analyses showing that depressive symptom severity predicts AF incidence.^15^ Notably, our depression classification used an integrative algorithm combining self-report, ICD-10 codes, and PHQ scores,^44^ whereas Lee et al. (2023) relied on a single-item measure.^15^ The consistency of findings across depression definitions supports the robustness of the association between AF and MDD. Our longitudinal analysis further emphasizes the importance of considering the timing of onset of comorbid AF and MDD. Future studies should explore age as a time-varying covariate, as it may significantly influence the temporal dynamics of the association between MDD and AF.

Mediation analyses further suggest that systemic biological pathways contribute to this bidirectional relationship. A high cardiovascular risk profile accounted for approximately one-third of the AF–MDD association, while inflammatory biomarkers accounted for a smaller but significant proportion. Specifically, inflammatory biomarkers (increased CRP, RF; decreased IGF-1) mediated 6.85% of the relationship, consistent with prior evidence linking systemic inflammation to both conditions.^20,21^ Importantly, these biomarkers primarily capture different facets of systemic inflammation, with CRP reflecting predominantly acute-phase inflammatory activity, while reduced IGF-1 and elevated RF may index more sustained or chronic inflammatory states. Inflammation has been shown to promote atrial remodeling by inducing cardiomyocyte apoptosis and fibrosis, which in turn alter atrial electrophysiology and facilitate the initiation and maintenance of AF.^21^ Inflammatory signals may disrupt glutamate neurotransmission, increase oxidative stress, and promote neurotoxicity, contributing to neural circuit dysfunction in MDD.^20^ Emerging evidence suggests that a subset of individuals with MDD may exhibit an inflammatory ‘biotype’, in which systemic inflammation amplifies motivational fluctuations, particularly anhedonia.^71^ This shared inflammatory pathway reinforces the bidirectional relationship between AF and MDD, suggesting that inflammation may act as a common underlying mechanism. Within this framework, AF may heighten vulnerability to depression, and conversely, depression may exacerbate AF—creating a vicious cycle characterized by dysregulated immune–brain–heart interactions. Although no current treatments specifically target inflammation in either condition, these findings point toward the potential for personalized anti-inflammatory therapies.^21,72^ Our findings also highlight the role of cardiovascular risk factors, including hypertension, dyslipidemia, hypertriglyceridemia, and obesity, which mediated 32.01% of the association between AF and MDD. However, as with all mediation analyses conducted in observational cohorts, these estimates do not establish causal pathways and may be influenced by unmeasured or residual confounding factors. These factors are well-established contributors to the risk of developing AF.^73–75^ Conversely, AF may further elevate cardiovascular risk by increasing myocardial oxygen demand, reducing coronary perfusion during arrhythmias, and promoting a heightened inflammatory response that accelerates both atherosclerosis and endothelial dysfunction.^23^ In depression, unhealthy behaviors such as smoking, physical inactivity, poor diet, and non-adherence to medications further contribute to an increased risk of cardiometabolic syndrome and coronary artery disease.^76^ These factors underscore that the cardiovascular risk pathway likely reflects a combination of biological and behavioral processes rather than a discrete mechanistic mediator. Beyond these behavioral factors, depression independently elevates the risk of developing coronary artery disease through inflammatory processes that may promote atherosclerosis.^6,19^ Importantly, our results align with prior UK Biobank research indicating that inflammation and metabolic syndrome mediate the association between depression and vascular dysfunction.^44^ Together, these findings underscore the need for integrated approaches that evaluate and target systemic inflammation and cardiovascular risk factors.

Our neuroimaging findings provide additional insight into potential neural substrates of AF–MDD comorbidity. Structurally, AF was associated with greater gray matter volume in key CAN hubs, including the ventromedial prefrontal cortex and insula, whereas MDD was associated with reductions in these regions. Functional analyses revealed parallel patterns, with increased CAN connectivity in AF and decreased connectivity in MDD. These findings are consistent with prior work demonstrating insular and prefrontal abnormalities in depression^25,77^ and emerging evidence of brain structural changes in AF.^33,78^ The absence of significant structural differences in the comorbid AF-MDD group suggests a non-linear interaction between these conditions, potentially reflecting compensatory or adaptive neurocardiac processes. Importantly, the magnitude of volumetric differences was small, and the cross-sectional imaging design precludes determination of temporal directionality or causal mechanisms. Nonetheless, these results highlight the CAN as a key neural interface linking central autonomic regulation with cardiovascular and affective processes (see Supplement for further discussion). Our findings also provide the first large-scale evidence of between-network connectivity differences: AF was associated with increased CAN–CEN, CAN–SN, and CAN–DMN coupling, whereas MDD and comorbid AF–MDD showed decreased or unchanged between-network connectivity. To our knowledge, this is the first study to systematically assess CAN-related connectivity patterns in comorbid AF and MDD. Finally, unlike earlier studies reporting amygdala volume alterations in MDD,^79^ we found no significant differences, which may reflect sample heterogeneity, including variation in depression subtypes, medication status, or the presence of remitted versus active symptoms.^80^

Exploratory analyses of peripheral autonomic signatures suggested that AF and AF–MDD conditions may reflect increased parasympathetic activity, consistent with prior literature in AF,^81,82^ despite contrasting with our initial hypothesis of sympathetic overactivity. However, heart rate variability metrics derived from ultra-short ECG segments during AF may largely reflect irregular R–R intervals rather than true vagal modulation, warranting cautious interpretation. The absence of heart rate variability differences in MDD contrasts with studies reporting reduced heart rate variability in depression,^83,84^ and may relate to methodological factors, including the use of ultra-short ECG recordings—although recent validation in the UK Biobank^56^ supports their comparability to standard indices—as well as the heterogeneous, population-based definition of depression, potentially diluting observable heart rate variability effects. Overall, these exploratory findings suggest differential peripheral autonomic signatures across groups and highlight the need for more granular autonomic assessments in future studies of AF–MDD comorbidity.

From a clinical perspective, these findings support the view that AF and depression share common systemic risk pathways that extend beyond traditional disease boundaries. The strong bidirectional associations observed in this study suggest that depression may represent both a risk marker and a potential modifier of AF progression, while AF itself may contribute to subsequent vulnerability to depressive illness. These observations underscore the importance of integrated risk assessment strategies that consider psychological health, cardiometabolic risk factors, and systemic inflammation in patients with AF. Conversely, individuals with depression may represent an underrecognized population at elevated risk for incident AF, highlighting the need for greater cardiovascular risk surveillance in psychiatric settings.

This study has several limitations. First, neuroimaging analyses were based on cross-sectional MRI data, precluding causal inference regarding the temporal ordering of brain alterations, AF, and MDD. Longitudinal neuroimaging studies are needed to determine whether structural and functional changes within the central autonomic network precede the onset of AF or depression, emerge as a consequence of disease progression, or reflect adaptive or compensatory mechanisms associated with comorbidity. Second, although mediation analyses highlighted a role for inflammatory pathways, the inflammatory markers available in the UK Biobank capture only a subset of systemic inflammatory processes. In particular, well-established markers of chronic low-grade inflammation in MDD were not available,^20^ limiting our ability to fully disentangle acute versus chronic inflammatory contributions. Future studies incorporating more comprehensive immunophenotyping may help refine the mechanistic understanding of immune–brain–heart interactions underlying AF–MDD comorbidity. Finally, depression was defined using a combination of self-report, hospital records, and brief symptom screening (PHQ-2), which may not fully capture the clinical heterogeneity of depressive disorders. While extensive sensitivity analyses were conducted, studies using in-depth clinical assessments and repeated symptom measurements would be valuable to further clarify the role of symptom severity, chronicity, and specific depressive dimensions in relation to AF.

## CONCLUSION

In this UK Biobank analysis, AF and MDD were robustly and bidirectionally associated, with temporal sequencing in both directions and partial statistical accounting by cardiovascular risk and inflammatory biomarkers. Across neurocardiac measures, AF and MDD showed opposing central autonomic network (CAN) structural and connectivity patterns, while comorbid AF-MDD exhibited distinct, non-additive profiles—supporting a systems-level model in which shared systemic risk intersects with disorder-specific neurocardiac adaptations. Although observational and cross-sectional components preclude direct causal inference, these findings motivate longitudinal and interventional studies to test whether targeting cardiometabolic and inflammatory risk, alongside integrated cardiovascular-mental health care, can reduce AF-MDD comorbidity and improve outcomes.

## Author Contributions

Concept and design: Verdonk, Hakimi, Khalsa. Processing of data: Verdonk, Hakimi, Misaki, Steinhäuser. Statistical analysis of data: Verdonk, Hakimi. Interpretation of data: all authors. Drafting of the manuscript: Verdonk, Talishinsky, Khalsa. Critical revision of the manuscript for important intellectual content: all authors. Obtained funding: Paulus, Khalsa. Supervision: Khalsa.

## Data Availability

Individual-level data used in the present study are available upon application to the UK Biobank. All other relevant data are available upon reasonable request to the authors.

https://www.ukbiobank.ac.uk

