## Supplemental file for "Shared Neurocardiac Pathways Linking Atrial Fibrillation and Depression: A UK Biobank Analysis"

**Competing Interests.**
CV, AT and JLS have no competing interests to disclose. SSK has performed scientific consulting for Jannsen Pharmaceuticals and served on a Data Safety Monitoring Board for Engrail Pharmaceuticals. OAA has received honoraria from J&J Medtech/Biosense Webster, Biotronik, Boston Scientific, Abbott, and Medtronic; holds stock in Anumana, NeuCures, and nference; and is a cofounder of Neufera. OAA also has a pending patent on Aorticorenal ganglion for neuromodulation.

**Table of contents**

**Supplementary Methods 1.** Sample size across data types and study groups……..……..Page 3
**Supplementary Methods 2.** MRI data processing……………………………………..…...Page 4
**Supplementary Methods 3.** Definition of cardiovascular risk profile.…………..……….....Page 7
**Supplementary Methods 4.** Computation of usHRV metrics……………………………..….Page 8
**Supplementary Methods 5.** Mediation analysis……………………..……..……..……….Page 9
**Supplementary Methods 6.** Survival analysis……...………………....………...…..……...Page 10
**Supplementary Results 1.** Sensitivity analyses………..………..…..…………..………...Page 11
**Supplementary Results 2.** Mediation findings...……………………..…………..………...Page 13

**Supplementary Methods 1.** Sample size across data types and study groups

The approach implemented in our study resulted in datasets with varying sample sizes depending on data availability for each data type (Table S1). This variability arises from differences in data collection protocols, missing data, and participant exclusions due to corrupted data files during processing, all of which are common to the UK Biobank dataset. The table below summarizes the number of participants in each study group—individuals with atrial fibrillation (AF), major depressive disorder (MDD), comorbid AF and MDD, and healthy comparisons (HCs)—across different data types, including neuroimaging data, electrocardiogram (ECG) data, inflammatory biomarkers, and cardiovascular (CV) risk factors.

**Table S1 -** Sample sizes by study groups and data type.

| | Groups | | | | Total |
|---|---|---|---|---|---|
| **Data type** | **HCs** | **AF+/MDD-** | **AF-/MDD+** | **AF+/MDD+** | |
| Structural MRI | 35,737 | 1,820 | 5,342 | 221 | 43,120 |
| Connectivity MRI | 33,948 | 1,716 | 5,793 | 243 | 41,700 |
| ECG data | 38,972 | 1,943 | 5,793 | 243 | 46,951 |
| Inflammation data | | | | | |
| CRP | 380,698 | 30,223 | 52,172 | 5,168 | 468,261 |
| IGF-1 | 379,471 | 30,121 | 51,993 | 5,158 | 466,743 |
| PR | 33,041 | 3,178 | 4,550 | 520 | 41,289 |
| CV risk factors | 391,227 | 31,224 | 72,909 | 6,817 | 502,177 |

AF: Atrial Fibrillation; MDD: Major Depressive Disorder; HCs: healthy comparisons.

**Supplementary Methods 2.** MRI data processing

Connectivity measures and graph theory metrics were computed for four large-scale brain networks: the central autonomic network (CAN), central executive network (CEN), salience network (SN), and default mode network (DMN). Table S2 lists the Brainnetome atlas regions corresponding to each network.[1] Supplementary Figure S1 shows a graphical summary of the regions of interest corresponding to each network.

**Table S2 -** Brainnetome atlas regions corresponding to large-scale brain networks examined in the present study.

| Brain region | Brainnetome atlas-based indexes | Functional networks |
|---|---|---|
| Left Mid cingulate cortex | 183 | Central Autonomic Network |
| Right Mid cingulate cortex | 184 | Central Autonomic Network |
| Left Anterior (agranular) insula | 165, 167 | Central Autonomic Network & Salience Network |
| Right Anterior (agranular) insula | 166, 168 | Central Autonomic Network & Salience Network |
| Left Mid (dysgranular) insula | 169, 173 | Central Autonomic Network & Salience Network |
| Right Mid (dysgranular) insula | 170, 174 | Central Autonomic Network & Salience Network |
| Left Posterior (granular) insula | 163, 171 | Central Autonomic Network & Salience Network |
| Right Posterior (granular) insula | 164, 172 | Central Autonomic Network & Salience Network |
| Left Amygdala | 211, 213 | Central Autonomic Network |
| Right Amygdala | 212, 214 | Central Autonomic Network |
| Left Thalamus | 231,233,235,237,239,241,243, 245 | Central Autonomic Network |
| Right Thalamus | 232,234,236,238,240,242,244, 246 | Central Autonomic Network |
| Left Anterior cingulate cortex | 177,179, 187 | Central Executive Network & Salience Network |
| Right Anterior cingulate cortex | 178,180, 188 | Central Executive Network & Salience Network |
| Left Dorsolateral prefrontal cortex | 3,15,17,23, 25 | Central Executive Network |
| Right Dorsolateral prefrontal cortex | 4,16,18,24, 26 | Central Executive Network |
| Left Middle temporal gyrus + anterior temporal cortex | 81,83,87, 95 | Default Mode Network |
| Right Middle temporal gyrus + anterior temporal cortex | 82,84,88, 96 | Default Mode Network |
| Left Angular gyrus / posterior parietal cortex | 143 | Default Mode Network |
| Right Angular gyrus / posterior parietal cortex | 144 | Default Mode Network |
| Left Dorsomedial prefrontal cortex | 1, 11 | Default Mode Network |
| Right Dorsomedial prefrontal cortex | 2, 12 | Default Mode Network |
| Left Ventromedial prefrontal cortex | 41,45,47, 49 | Default Mode Network & Central Autonomic Network |
| Right Ventromedial prefrontal cortex | 42,46,48, 50 | Default Mode Network & Central Autonomic Network |
| Left Posterior cingulate cortex | 175, 181 | Default Mode Network |
| Right Posterior cingulate cortex | 176, 182 | Default Mode Network |
| Left Hippocampus | 215, 217 | Default Mode Network |
| Right Hippocampus | 216, 218 | Default Mode Network |

**Figure S1.** Graphical summary of the regions of interest corresponding to each network.

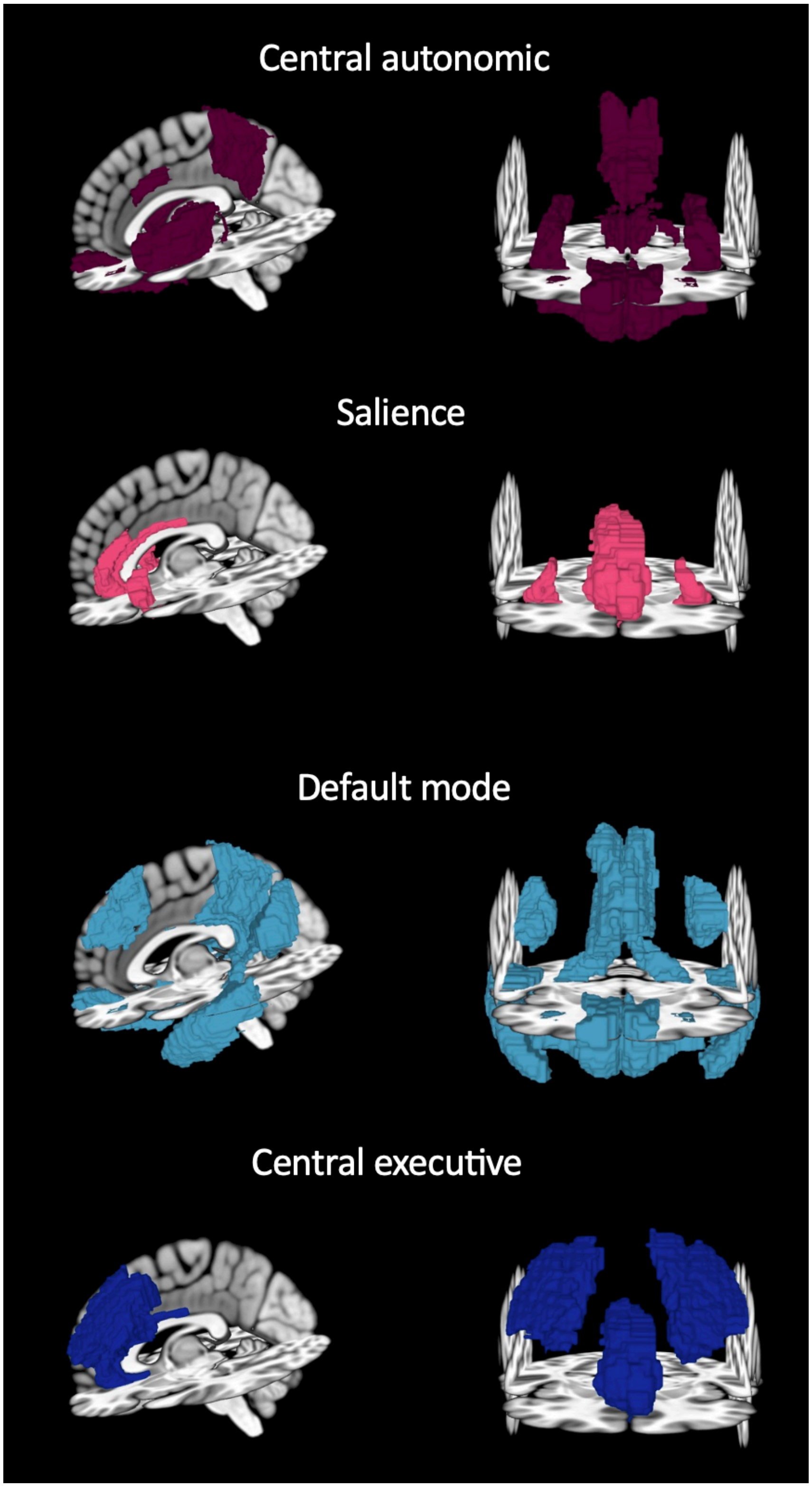

**Supplementary Methods 3.** Definition of cardiovascular risk profile

The cardiovascular (CV) risk profile was defined as ‘high’ if presence of 3 or more of the following CV risk factors: unhealthy waist circumference, hypertension, dyslipidemia, hypertriglyceridemia, and hyperglycemia.[2] These measures were assessed during the initial visit of enrollment. Waist circumference and blood pressure were recorded by a trained nurse. Waist circumference was measured at the narrowest part of the torso using a 200-cm SECA tape measure. Unhealthy waist circumference was defined based on established thresholds: ≥88 cm for women and ≥102 cm for men.[3] Blood pressure measurements were taken twice using the IntelliSense blood pressure monitor (model HEM-907XL, Omron) after participants had rested for at least 5 minutes.[4] The average of the two readings was used to define hypertension, which was categorized as a systolic blood pressure of 140 mmHg or higher, a diastolic blood pressure of 90 mmHg or higher, or a hypertension diagnosis utilizing the following ICD-10 codes: I10 (essential hypertension; UK Biobank field: 131286), I15 (secondary hypertension; UK Biobank field: 131294), I13 (hypertensive heart and renal disease; UK Biobank field: 131292), and O10 (pre-existing hypertension complicating pregnancy, childbirth, and the puerperium; UK Biobank field: 132180). Dyslipidemia was defined as a high-density lipoprotein cholesterol level below 40 mg/dL for men or below 50 mg/dL for women (to convert to millimoles per liter, multiply by 0.0259). Hypertriglyceridemia was classified as triglyceride levels of 150 mg/dL or higher (to convert to millimoles per liter, multiply by 0.0113). Hyperglycemia was identified as fasting blood glucose levels exceeding 110 mg/dL (to convert to millimoles per liter, multiply by 0.0555).

**Supplementary Methods 4.** Computation of usHRV metrics

Ultra-short heart rate variability (usHRV) metrics were extracted from 15-second resting electrocardiogram (ECG) recordings using a processing pipeline implemented in MATLAB 2020b (MathWorks®). The pipeline relied on the HRVAS-master toolbox (version 1.0.2),[5] scripts developed by Candia-Rivera and publicly available on GitHub (https://github.com/diegocandiar),[6] additional custom scripts, and followed a previously described methodology.[7]
ECG recordings were retrieved from the UK Biobank dataset. The initial dataset comprised 54,980 ECG recordings, of which 8,003 were identified as corrupted during preprocessing and excluded, resulting in 46,967 recordings retained for usHRV computation.
R-peak detection was performed using a modified Pan–Tompkins algorithm,[8] as implemented by Sedghamiz (2014) and available via MathWorks website. Interbeat intervals (IBIs) were computed from detected R-peaks and used for usHRV feature extraction. The Root Mean Square of Successive Differences (RMSSD) was computed using the *timeDomainHRV()* function from the HRVAS toolbox, while the Standard Deviation of Successive Differences (SDSD) was calculated using custom scripts. Successive differences were defined as the variation between consecutive IBIs, and both RMSSD and SDSD are primarily proposed to reflect short-term autonomic modulation of heart rate.[7,9,10] High-Frequency Power (PHF), representing power in the 0.15–0.40 Hz spectral band, was computed using a power spectral density approach. Specifically, PHF was derived as the median normalized high-frequency metric from the *compute_PWVD()* function developed by Candia-Rivera and available on GitHub (https://github.com/diegocandiar) [6]. PHF is considered a marker of parasympathetic activity.[9,10]
Importantly, these ultra-short HRV (usHRV) metrics from the UK Biobank dataset have been validated through their strong agreement with standard HRV measures obtained from 6-minute ECG recordings.[7]

**Supplementary Methods 5.** Mediation analysis

To examine whether the association between AF and MDD was mediated by a high CV risk profile, we implemented a single mediator model. First, we regressed the CV risk profile on AF status, denoting the corresponding regression coefficient as ***a***. Next, we regressed MDD status on both the CV risk profile and AF status, where the regression coefficients were ***b*** for the CV risk profile and ***c'*** for AF status. The indirect effect of AF on MDD via the CV risk profile was calculated as ***a********b***, with its confidence interval estimated using a bootstrapping approach with 500 replications.[11] The proportion of the total effect mediated by the CV risk profile was computed as the ratio of the indirect effect to the total effect: (***a********b***)/(***c'***+(***a********b***)), quantifying the extent to which the CV risk profile explains the observed association between AF and MDD (Figure S2A).
To test whether the association between AF and MDD was mediated by inflammatory response, assessed through markers such as C-reactive protein (CRP), insulin-like growth factor (IGF-1), and rheumatoid factor (RF), we implemented a multiple mediator modeling as follows. For each mediator, we computed the mediated effect as the coefficient labelled $\boldsymbol{a}_k$, $k$ [1…*M*] (*M* being the number of mediators) in regressing the mediator on the AF condition status. Similarly, we computed the indirected effect as the coefficient labelled $\boldsymbol{b}_k$, $k$ [1…*M*] (*M* being the number of mediators)in regression the MDD condition status on the mediator. The indirect effect was computed as the sum of the product $\boldsymbol{a}_k * \boldsymbol{b}_k$ and its confidence interval and its magnitude were computed similarly to the single mediator model (Figure S2B).

**Figure S2** - Graphical representation of the single mediator model **(A)** and the multiple mediator model **(B)** used in the present study.

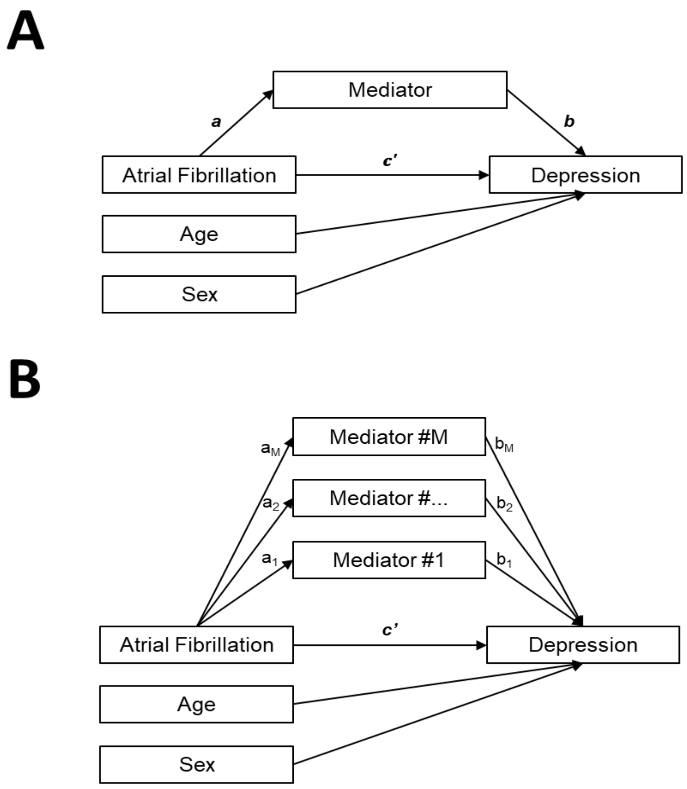

**Supplementary Methods 6.** Survival analysis

**Figure S3 - Design of survival analyses examining incident diagnoses of Major Depressive Disorder (MDD) and Atrial Fibrillation (AF)**. **(A)** Incident MDD diagnosis: For participants diagnosed with AF who later developed MDD (AF+/MDD+), follow-up began at the date of AF diagnosis and ended at the date of MDD diagnosis. For control participants without AF (AF-/MDD+ and AF-/MDD-), follow-up started at an age-matched AF diagnosis date from AF+/MDD+ participants. For AF-/MDD+ individuals, follow-up ended at the date of MDD diagnosis; for AF-/MDD- participants, data were censored on August 1, 2022—the date of the most recent recorded MDD diagnosis. Participants with AF who did not develop MDD (AF+/MDD-) were followed from their AF diagnosis date until August 1, 2022. **(B)** Incident AF diagnosis: For participants diagnosed with MDD who later developed AF (MDD+/AF+), follow-up began at the date of MDD diagnosis and ended at the date of AF diagnosis. For control participants without MDD (MDD-/AF+ and MDD-/AF-), follow-up started at an age-matched MDD diagnosis date from MDD+/AF+ participants. MDD-/AF+ individuals were followed until the date of AF diagnosis, while MDD-/AF- participants were censored on July 1, 2022—the date of the most recent recorded AF diagnosis. MDD+/AF- participants were followed from their MDD diagnosis date until July 1, 2022.

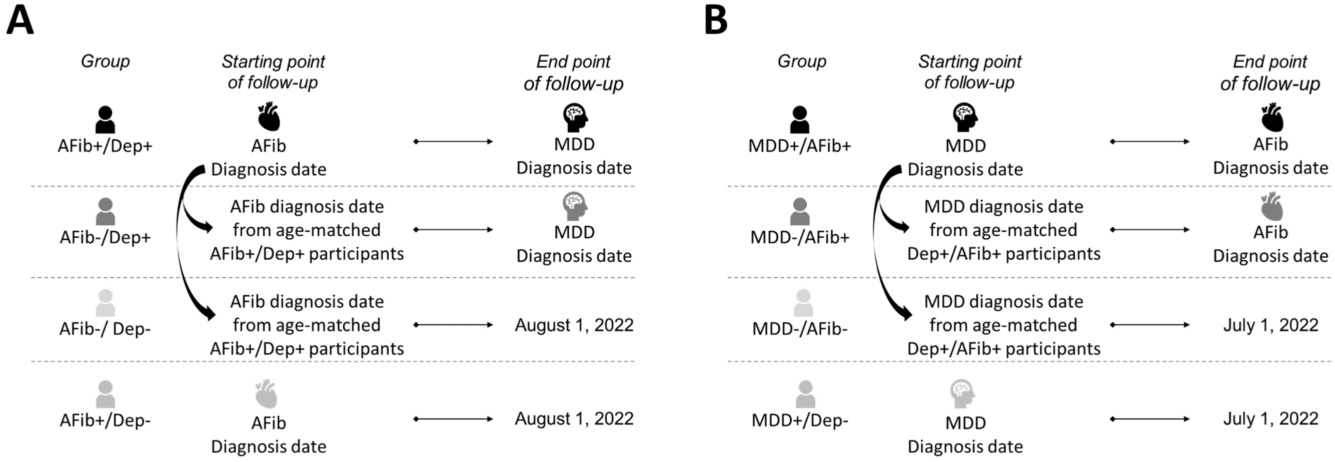

**Supplementary Results 1.** Sensitivity analyses

To assess the robustness of the association between AF and MDD across different levels of depressive symptom severity, we conducted a series of sensitivity analyses using alternative PHQ-2–based case definitions. In addition to the primary definition (PHQ-2 total score ≥ 3), we examined two symptom-focused subgroups: (i) severe depressive symptoms, defined by endorsement of “Nearly every day” on at least one of the two PHQ-2 items, and (ii) low depressive symptoms, defined by endorsement of “Several days” on at least one PHQ-2 item.

1. **Severe MDD Analysis**

    a. **Descriptive statistics**

Among UK Biobank participants with complete PHQ-2 item-level data at baseline, 15,504 participants (3.54%) met criteria for severe depressive symptoms according to this definition. Within this subset, 1,235 participants (8%) exhibited comorbid AF and severe depressive symptoms.

    b. **Results**

Consistent with the primary analysis, AF and depressive symptoms remained significantly and bidirectionally associated when restricting the definition of depression to severe PHQ-2 symptomatology. Participants with AF were more likely to have MDD with severe depressive symptoms than those without AF ($\beta=0.26$; $p<0.001$; OR=1.29, 95% CI [1.21; 1.37]). Conversely, individuals with MDD characterized by severe depressive symptoms had a higher likelihood of AF compared with those without MDD ($\beta=0.31$; $p<0.001$; OR=1.37, 95% CI [1.28; 1.45]).

Notably, effect sizes in this severe-symptom analysis were attenuated relative to those observed using the broader MDD definition (OR=1.37 and 1.41 in the primary analyses, respectively), suggesting that the strength of the AF–MDD association decreases as depressive symptom severity increases.

2. **Low MDD Analysis**

To further examine whether this pattern reflected a severity-dependent gradient, we conducted an additional sensitivity analysis focusing on participants with lower-severity depressive symptoms, defined by endorsement of “Several days” on at least one PHQ-2 item.

    a. **Descriptive statistics**

Among UK Biobank participants with complete PHQ-2 item-level data at baseline, 33,634 participants (7.38%) met criteria for low depressive symptoms according to this definition. Within this subset, 2,755 participants (8.19%) exhibited comorbid AF and low depressive symptoms.

    b. **Results**

In this subgroup, the bidirectional association between AF and depression remained robust. Participants with AF showed an increased likelihood of having MDD with low depressive symptoms than those without AF ($\beta=0.33$; $p<0.001$; OR=1.39, 95% CI [1.33; 1.45]), and individuals with MDD characterized by low depressive symptoms had a higher likelihood of AF ($\beta=0.37$; $p<0.001$; OR=1.45, 95% CI [1.39; 1.51]).

Effect sizes in this analysis were comparable to, or slightly stronger than, those observed in the primary analysis.

### 3. Continuous severity analysis within the depressed subgroup

Taken together, the categorical sensitivity analyses based on severe and low depressive symptom definitions indicated that the AF-depression association remained robust across symptom severity levels, while suggesting a relative attenuation of effect sizes in more severe symptom groups. To clarify whether this pattern reflected a true inverse relationship between depressive symptom severity and AF comorbidity, we conducted an additional sensitivity analysis restricted to participants with depression, modeling depressive symptom severity as a continuous variable.

In this analysis, AF status was modeled as the outcome, and depressive symptom severity was entered as a continuous predictor using the PHQ-2 total score.
Within this depressed subgroup, PHQ-2 severity was not significantly associated with AF status ($\beta=-0.009$; $p=0.23$). The direction of the effect was negative but small in magnitude and statistically non-significant, providing no evidence for an inverse relationship between depressive symptom severity and AF comorbidity.

Together with the categorical sensitivity analyses, these results indicate that while the AF–depression association is robust across different symptom-based definitions of depression, variation in depressive symptom severity among individuals with depression does not appear to meaningfully influence AF comorbidity risk. This suggests that the observed attenuation of effect sizes in more severe symptom groups likely reflects differences in case definition and sample composition rather than a monotonic severity-dependent relationship.

**Supplementary Results 2.** Mediation findings

**Table S3. Statistical results from the mediation analysis**. Total effect (***c***), Mediated effect (***a***), Direct effects (***b*** and ***c'***), and Indirect effect in the association between Atrial Fibrillation and Major Depressive Disorder through the examined mediators in the present study.

| **Mediator** | **Statistical results** | |
|---|---|---|
| | **β [95% CI]** | **P-value** |
| **CV risk profile** | | |
| Total effect (***c***) | 0.360 [0.331;0.388] | <0.001 |
| Mediated effect (***a***) | 0.482 [0.455;0.508] | <0.001 |
| Direct effects (***b***) | 0.326 [0.302; 0.349] | <0.001 |
| Direct effects (***c'***) | 0.333 [0.304;0.361] | <0.001 |
| Indirect effect | 0.157 [0.146; 0.169] | - |
| **Inflammatory** | | |
| Total effect (***c***) | 0.334 [0.241;0.426] | <0.001 |
| Mediated effect | | |
| $\boldsymbol{a_1}$ | 0.679 [0.496;0.863] | <0.001 |
| $\boldsymbol{a_2}$ | 0.676 [-0.016;1.367] | 0.06 |
| $\boldsymbol{a_3}$ | -0.368 [-0.563;-0.174] | 0.001 |
| Direct effects (***b***) | | |
| $\boldsymbol{b_1}$ | 0.018 [0.014; 0.023] | <0.001 |
| $\boldsymbol{b_2}$ | 0.001 [0.304;0.361] | 0.21 |
| $\boldsymbol{b_3}$ | -0.027 [-0.032;-0.022] | <0.001 |
| Direct effects (***c'***) | | |
| $\boldsymbol{c'_1}$ | 0.321 [0.227;0.413] | <0.001 |
| $\boldsymbol{c'_2}$ | 0.334 [0.240;0.426] | <0.001 |
| $\boldsymbol{c'_3}$ | 0.323 [0.229;0.415] | <0.001 |
| Indirect effect | 0.157 [0.146; 0.169] | - |

95% CI: 95% Confident interval; CV risk profile: Cardio-vascular risk profile;
